# Dose-dependent effect of nuts on blood pressure: a systematic review and meta-analysis of randomized controlled trials

**DOI:** 10.1101/2021.05.21.21257564

**Authors:** Ahmad Jayedi, Tauseef Ahmad Khan, Amin Mirrafiei, Bahareh Jabbarzadeh, Yasaman Hosseini, Sheida Motlagh, Aliyu Tijani Jibril, Hossein Shahinfar, Sakineh Shab-Bidar

**Author notes:** **Correspondence to:** Sakineh Shab-Bidar, Associate Professor, Department of Community Nutrition, School of Nutritional Sciences and Dietetics, Tehran University of Medical Sciences, P. O. Box 14155/6117, Tehran, Iran Telefax.

## Abstract

**Objective:** Traditional pairwise meta-analyses indicated that nuts consumption can improve blood pressure. We iamed to determine the dose-dependent effect of nuts on systolic (SBP) and diastolic blood pressure (DBP) in adults.

**Methods:** A systematic search was undertaken in PubMed, Scopus, and ISI Web of Science till March 2021. Randomized controlled trials (RCT) evaluating the effects of nuts on SBP and DBP in adults were included. We estimated change in blood pressure for each 20 g/d increment in nut consumption in each trial and then, calculated mean difference (MD) and 95%CI using a random-effects model. We estimated dose-dependent effect using a dose-response meta-analysis of differences in means. The certainty of evidence was rated using the GRADE instrument, with the minimal clinically important difference being considered 2 mmHg.

**Results:** A total of 31 RCTs with 2784 participants were included. Each 20 g/d increase in nut consumption reduced SBP (MD: -0.50 mmHg, 95%CI: -0.79, -0.21; I^2^ = 12%, n = 31; GRADE = moderate certainty) and DBP (MD: -0.23 mmHg, 95%CI: -0.38, -0.08; I^2^ = 0%, n = 31; GRADE = moderate certainty). The effect of nuts on SBP was more evident in patients with type 2 diabetes (MD: -1.31, 95%CI: -2.55, -0.05; I^2^ = 31%, n = 6). The results were robust in the subgroup of trials with low risk of bias. Levels of SBP decreased proportionally with the increase in nuts consumption up to 40 g/d (MD_40g/d_: -1.60, 95%CI: -2.63, -0.58), and then appeared to plateau with a slight upward curve. A linear dose-dependent reduction was seen for DBP, with the greatest reduction at 80 g/d (MD_80g/d_: -0.80, 95%CI: -1.55, -0.04).

**Conclusions:** The available evidence provides a good indication that nut consumption can result in a small improvement in blood pressure in adults. Well-designed trials are needed to confirm the findings in long term follow-up.

## Introduction

High blood pressure is the leading risk factor for cardiovascular disease (CVD) and death across the globe. In 2019, elevated blood pressure attributed for about 10.8 million death in the world, accounting for 19% of all deaths (1). Despite effective antihypertensive medications (2), the global burden of high blood pressure is still high (3).

The effect of eating habits on levels of blood pressure have long been investigated. There is evidence that unhealthy eating habits such as high sodium and low potassium intake, alcohol drinking, and poor diet quality may contribute to developing hypertension (4). Adiposity is another diet-related condition that is highly associated with the risk of developing hypertension (5). Epidemiologic research has also suggested that high nut consumption was associated with a lower risk of developing hypertension (6).

Nuts are rich in mono- and polyunsaturated fatty acids, minerals such as magnesium and potassium, fibers, and antioxidants and thereby can confer protection against hypertension (7). Several meta-analyses of randomized controlled trials (RCT) have therefore been undertaken of the effects of nuts on blood pressure levels and have indicated that increasing the consumption of nuts can significantly decrease blood pressure levels (8-11).

However, existing meta-analyses only performed pairwise comparisons between intervention and control groups and did not consider differences between doses of nuts consumption in intervention groups across trials, as well as between intervention and control groups in each trial. The identification of dose-response associations is an important part of the analyses in nutrition research that can help determine whether the increasing the level of dose is effective, and to select the optimal dose for implementing the most effective interventions (12, 13).

Dose-response meta-analysis of differences in means is a new statistical approach that helps to address these considerations when evaluating the effects of a specific intervention on continuous outcomes (14). Dose-response meta-analysis of differences in means can present valuable information on the effects of different doses of an intervention on a continuous outcome that cannot be obtained by traditional pairwise meta-analyses and can determine the shape of dose-dependent effect. We, therefore, aimed to perform a systematic review and dose-response meta-analysis of RCTs to evaluate the effects of different doses of nuts consumption on systolic (SBP) and diastolic blood pressure (DBP) in adults.

## Methods

The present dose-response meta-analysis has been reported following the Preferred Reporting Items for Systematic Reviews and Meta-analyses: the PRISMA statement (15). The protocol of the systematic review was registered in PROSPERO (CRD42021247346).

### Systematic search

To find potential eligible RCTs for inclusion in this review, we searched PubMed, Scopus, and ISI Web of Science up to March 20201. We supplemented the database search by manually reviewing the reference lists of all existing related reviews. The search in the databases and reference lists was restricted to articles published in English. We combined keywords related to intervention, outcome, and study design to find potential eligible RCTs. The complete search strategy is described in **Supplementary Table 1**. Teams of two reviewers independently screened titles and abstracts according to the pre-defined inclusion and exclusion criteria to identify potential eligible trials.

### Eligibility criteria

We applied PICOS (population, intervention, comparator, outcome, and study design) framework to define our inclusion and exclusion criteria. Published human intervention studies were considered eligible for inclusion in the present meta-analysis if they had the following criteria: 1) RCTs, either with parallel or cross-over design, conducted in adults aged 18 years or older, regardless of health status; 2) evaluated the effect of nuts (mixed nuts or one of the nuts subtypes) on SBP and/or DBP; 3) compared the effect of different doses (g/d) of a specific nut on blood pressure across more than one study arm (*e*.*g*, 60 g/d vs 30 g/d pistachio) or compared the effect of the specific amount of nuts (g/d) against a nut-free diet; 4) considered the change in SBP and/or DBP as the primary or one of the secondary outcomes; 5) provided mean and standard deviation (SD) of change in DBP or SBP across study arms or reported sufficient information to estimate those values; and 6) reported the number of participants in each study arm.

Trials with non-randomized design, quasi-experimental studies, trials conducted in adolescents (under 18 years of age), pregnant and lactating women, those that applied an active control group with possible effects on blood pressure (*e*.*g*, nuts vs fruit or nuts vs omega-3) (16), trials that did not specify the amount of nuts consumption in the control group (*e*.*g*, control group was usual or healthy diet without reporting the amount of nuts consumption), as well as trials that compared the effects of two different types of nuts (*e*.*g*, pistachio vs almond) were excluded. We excluded trials that compared the effects of two different types of nuts because of their active control group.

### Data extraction

Two reviewers (HS and SM) independently and in duplicate screened the full texts of eligible trials and extracted the following data: author and year, population location, study design and duration, characteristics of the population (mean age +/- SD, baseline BMI, health status), total sample size, intervention characteristics (type and dose of nuts consumption), comparison group, calorie restriction, physical activity, behavioral support, outcome measures and main results for the outcomes included. Disagreements between the two reviewers were resolved by discussion.

### Risk of bias (quality) assessment

Two reviewers (AJ and SSB) independently and in duplicate performed risk of bias assessments using the Cochrane risk of bias tool (17). An overall quality score was given to the trials based on bias domains: good (≤1/5 items were unknown and none were high), fair (≤2/5 items were unclear or at least one high), and high risk of bias (≥2/5 items were high). Disagreements regarding the risk of bias assessment were resolved by discussion.

### Statistical analysis

We considered weighted mean difference (MD) and 95% confidence interval (CI) of change in SBP and DBP as the effect size for reporting the results of the present systematic review. First, we calculated changes from baseline blood pressure in each study arm. If the mean values and SDs of changes were not available, we calculated these values by using data from measures before and after the intervention, according to the guidelines of the Cochrane Handbook (18). When standard errors instead of SDs were presented, the former was converted to SDs (19). If studies reported medians and interquartile ranges, we used the median to impute the missing mean and calculated SDs by dividing interquartile ranges by 1.35 (19). If none of these options was available, we imputed the missing SDs using pooled SDs obtained from other trials included in our meta-analysis (20).

Second, we used the method introduced by Crippa and Orsini (14) to calculate MD and its corresponding SD of change in SBP and DBP for each 20 g/d increment in nuts consumption in the intervention group relative to the control group in each trial. This method requires dose (g/d) of nuts consumption in each study arm, the mean and its corresponding SD of change in SBP and DBP in each study arm, and the number of participants in each arm. Trial-specific results were pooled using a random-effects model (21).

We then performed a series of pre-defined subgroup analyses based on health status, baseline weight (normal weight or overweight/obese), presence of calorie restriction, physical activity, or behavioral support in the intervention program, as well as the type of funding (industrial vs university), follow-up duration, and risk of bias assessment. Influence analysis was carried out to test the potential impact of each trial on the pooled effect size. The potential for publication bias was tested using Egger’s test (22), Begg’s test (23), and by inspection of funnel plots. We assessed heterogeneity quantitatively using the I^2^ statistic and performed a χ^2^ test for homogeneity (P_heterogeneity_> 0.10) (24).

Finally, we performed a dose-response meta-analysis to clarify the shape of the effect of different doses of nuts on SBP and DBP (14). Statistical analyses were conducted using STATA software version 16.1. A two-tailed P value of less than 0.05 was considered significant.

### Grading the evidence

We assessed the certainty of the evidence using the GRADE approach (25). Minimal clinically important difference (MCID) for SBP and DBP was defined as 2 mmHg (26).

## Results

The database and reference lists search identified 4502 articles. We excluded 291 duplicates and further 4103 articles based on screening of the title and abstract. Eventually, 108 full texts were fully reviewed for eligibility. Of those, 31 randomised trials with 2784 participants were eligible for inclusion in dose-response meta-analysis (27-57). Detailed screening and data extraction processes are depicted in **Supplementary Figure 1**, and list of studies excluded based on reviewing the full texts are presented in **Supplementary Table 2**.

### Characteristics of randomised trials

**Supplementary Table 3** presents the general characteristics of the trials included in the meta-analysis. Eligible trials were published between 2007 and 2020. Twelve trials were conducted in individuals at high risk of CVD (with at least one cardiometabolic risk factor) (36, 37, 40, 42, 45-47, 49-51, 53, 55), six trials each in patients with type 2 diabetes (30, 38, 41, 43, 48, 56) and healthy individuals (27, 32-34, 50, 52), four in patients with the metabolic syndrome (29, 44, 54, 57), and three in those with a history of CVD (28, 31, 39). Fourteen trials were conducted exclusively in individuals with overweight/obesity (30-33, 35, 36, 40, 42, 45, 47, 49, 50, 52, 53), and the remainders in mixed populations (27-29, 34, 37-39, 41, 43, 44, 46, 48, 51, 54-57).

The dose of nut consumption ranged between 15 to 85 g/d. Three trials compared the effect of two doses of a specific type of nut against a nut free diet (52, 54, 55), and the remainder compared the effects of a nut-supplemented diet against a nut free diet (27-51, 53, 56, 57). One study compared the effect of two different types of nuts against a nut free diet and thus, was included as two study arms in the analysis (44).

Of the trials, 22 lasted ≤12 weeks (28-34, 38, 40-45, 47, 48, 51-55, 57), seven trials lasted >12 to ≤24 weeks (35, 37, 39, 46, 49, 50, 56), and two trials >24 weeks (27, 36). Six trials applied behavioral support (33, 35, 36, 42, 49, 50), five trials implemented a calorie-restricted diet (33, 36, 49, 50, 56), and three implemented exercise (36, 49, 50) alongside nuts supplementation. Of the trials, 12 were rated to have low risk of bias (27, 28, 32, 34, 40, 41, 45, 46, 49, 50, 53, 56), nine trials had moderate risk of bias (some concerns) (29-31, 35, 39, 42-44, 47), and 10 were rated to have high risk of bias (33, 36-38, 48, 51, 52, 54, 55, 57) (**Supplementary Table 4**).

### The effect of nuts on systolic blood pressure

All 31 trials reported data for the effect of nuts on SBP. Each 20 g/d increment in nuts consumption reduced SBP by -0.50 mmHg (95%CI: -0.79, -0.29; I^2^ = 12%, **Supplementary Figure 2**). The effect size remained significant after step-wise exclusion of each study from the main analysis (MD range: -0.43 to -0.57).

**Table 1** presents the results of the subgroup analyses. The results remained significant in healthy individuals, in patients with type 2 diabetes and those with overweight/obesity, trials with a follow-up ≤12 weeks, those with low risk of bias, and trials that received industrial funding, though tests for subgroup deference were not significant. There was an indication of a larger effect size in trials that implemented calorie restriction (MD: -1.25; 95%CI: -2.16, -0.24). There was no indication for publication bias with Egger’s test (P = 0.24), Begg’s test (P = 0.39), or with the funnel plot (**Supplementary Figure 3**).

**Table 1.** Subgroup analysis of the effect of nuts consumption (20 g/d increase) on systolic blood pressure.

|  | **n** | **Mean difference (95%CI)** | **I<sup>2</sup> (%), P<sub>heterogeneity</sub>** | **P<sub>between</sub>** |
| --- | --- | --- | --- | --- |
| **All trials** | 31 | -0.50 (-0.79, -0.21) | 12%, 0.28 | - |
| **Health status** |  |  |  | 0.45 |
| High risk | 12 | -0.36 (-0.69, -0.02) | 0%, 0.80 |  |
| Type 2 diabetes | 6 | -1.31 (-2.55, -0.05) | 31%, 0.20 |  |
| Healthy | 6 | -0.93 (-2.10, 0.24) | 48%, 0.09 |  |
| Metabolic syndrome | 4 | -0.42 (-1.10, 0.26) | 0%, 0.55 |  |
| CVD | 3 | -0.13 (-1.67, 1.40) | 0%, 0.55 |  |
| **Follow-up duration** |  |  |  | 0.65 |
| ≤12 weeks | 22 | -0.46 (-0.79, -0.13) | 3%, 0.42 |  |
| 12 to ≤24 weeks | 7 | -0.93 (-1.93, 0.08) | 28%, 0.22 |  |
| >24 weeks | 2 | -0.38 (-1.27, 0.50) | 0%, 0.57 |  |
| **Risk of bias** |  |  |  | 0.08 |
| Low | 12 | -0.92 (-1.47, -0.38) | 35%, 0.11 |  |
| Moderate | 9 | -0.02 (-0.52, 0.49) | 0%, 0.53 |  |
| High | 10 | -0.35 (-0.86, 0.16) | 0%, 0.80 |  |
| **Weight status** |  |  |  | 0.73 |
| Normal weight | 25 | -0.56 (-0.95, -0.18) | 2%, 0.43 |  |
| Overweight/obese | 6 | -0.39 (-0.88, 0.11) | 8%, 0.37 |  |
| **Calorie restriction** |  |  |  | 0.09 |
| Yes | 5 | -1.20 (-2.16, -0.24) | 11%, 0.34 |  |
| No | 26 | -0.41 (-0.70, -0.13) | 0%, 0.53 |  |
| **Physical activity** |  |  |  | 0.16 |
| Yes | 3 | -1.32 (-3.25, 0.61) | 54%, 0.11 |  |
| No | 28 | -0.43 (-0.70, -0.17) | 0%, 0.59 |  |
| **Behavioral support** |  |  |  | 0.29 |
| Yes | 6 | -0.95 (-1.94, 0.05) | 8%, 0.37 |  |
| No | 25 | -0.45 (-0.75, -0.15) | 2%, 0.43 |  |
| **Funding** |  |  |  | 0.51 |
| Industrial | 19 | -0.49 (-0.74, -0.23) | 0%, 0.91 |  |
| University | 12 | -0.53 (-1.74, 0.69) | 44%, 0.05 |  |
| **Abbreviations:** CVD, cardiovascular disease. |  |  |  |  |

Dose-dependent effects of nuts on levels of SBP are indicated in **Figure 1**, upper panel and **Table 2**. Levels of SBP decreased proportionally with the increase in nuts consumption up to 40 g/d (MD_40g/d_: -1.60, 95%CI: -2.63, -0.58), and then appeared to plateau with a slight upward curve (P_nonlinearity_ = 0.08, P_dose-response_= 0.002).

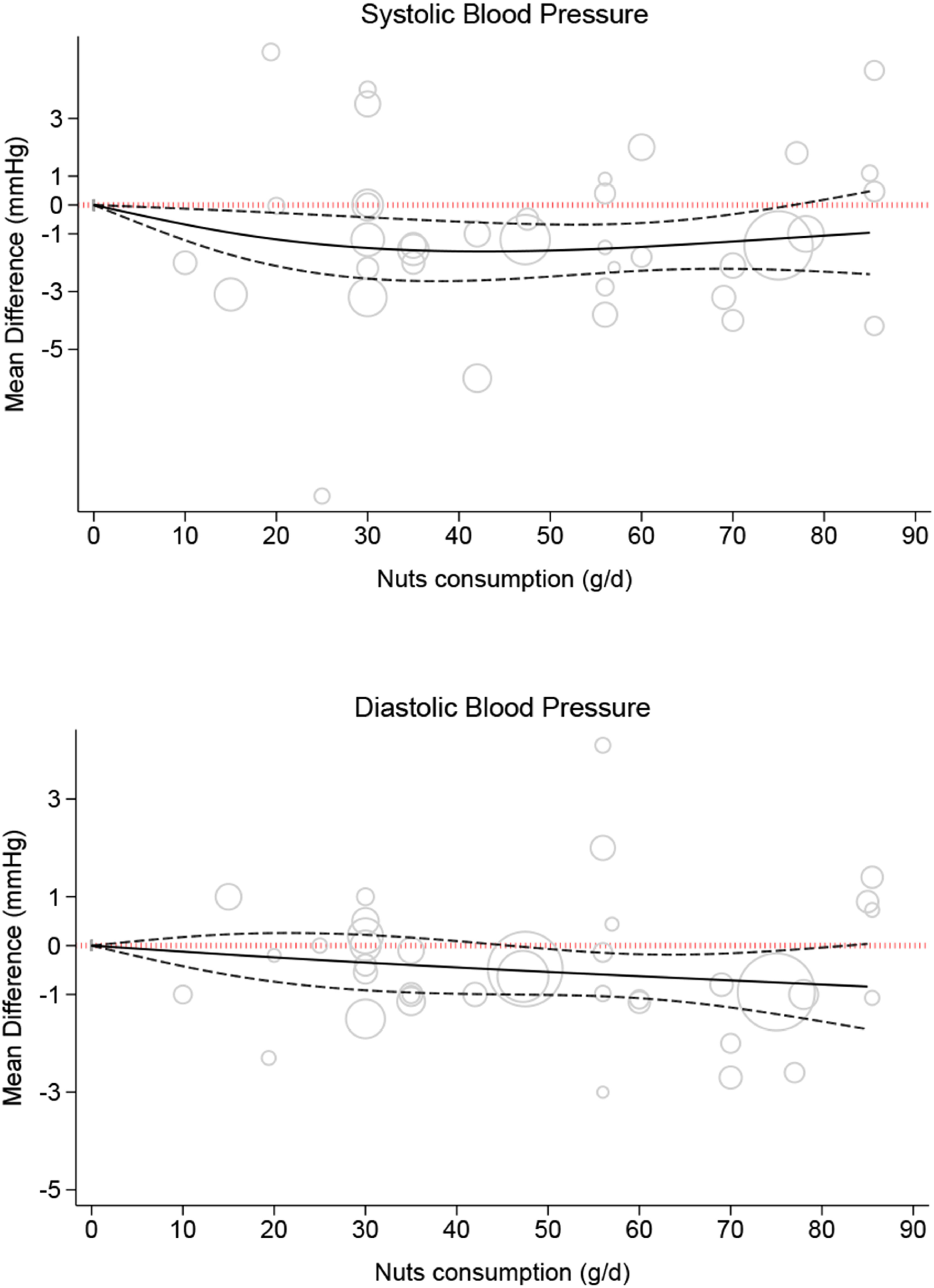

**Table 2.** The effects of different doses of nuts on blood pressure form the nonlinear dose-response meta-analysis (mean difference and 95% confidence interval).

| Nuts intake<br>(g/d) | 0<br>(Ref) | 10 | 20 | 30 | 40 | 50 | 60 | 70 | 80 |
| --- | --- | --- | --- | --- | --- | --- | --- | --- | --- |
| SBP (mmHg) | 0 | -0.68<br>(-1.22, -0.13) | -1.19<br>(-2.11, -0.27) | -1.49<br>(-2.55, -0.43) | -1.60<br>(-2.63, -0.58) | -1.58<br>(-2.48, -0.67) | -1.45<br>(-2.28, -0.62) | -1.27<br>(-2.21, -0.32) | -1.06<br>(-2.31, 0.18) |
| DBP (mmHg) | 0 | -0.12<br>(-0.42, 0.17) | -0.24<br>(-0.76, 0.26) | -0.35<br>(-0.91, 0.22) | -0.45<br>(-0.99, 0.09) | -0.54<br>(-1.01, -0.07) | -0.63<br>(-1.08, -0.18) | -0.71<br>(-1.27, -0.16) | -0.80<br>(-1.55, -0.04) |

**Abbreviations:** DBP, diastolic blood pressure; SBP, systolic blood pressure.

### The effect of nuts on diastolic blood pressure

Each 20 g/d increment in nuts consumption significantly reduced DBP (MD: -0.23; 95%CI: - 0.38, -0.08; I^2^ = 0%, **Supplementary Figure 4**). Sensitivity analysis did not show evidence of a heavy impact of each individual trial on pooled effect size (MD range: -0.21 to -0.25). **Table 3** indicates the subgroup analyses. The effect size was significant in patients at high risk of CVD and those with overweight/obesity, short-term follow-up trials, and in trials with low risk of bias. There was no indication for publication bias (Egger’s test P = 0.72, Begg’s test P = 0.71; **Supplementary Figure** 5).

**Table 3.** Subgroup analysis of the effect of nuts consumption (20 g/d increase) on diastolic blood pressure.

|  | **n** | **Mean difference (95%CI)** | **I<sup>2</sup> (%), P<sub>heterogeneity</sub>** | **P<sub>between</sub>** |
| --- | --- | --- | --- | --- |
| **All trials** | 31 | -0.23 (-0.38, -0.08) | 0%, 0.98 | - |
| **Health status** |  |  |  | 0.89 |
| High risk | 12 | -0.22 (-0.41, -0.04) | 0%, 0.88 |  |
| Type 2 diabetes | 6 | -0.26 (-0.92, 0.39) | 0%, 0.88 |  |
| Healthy | 6 | -0.32 (-0.78, 0.13) | 0%, 0.96 |  |
| Metabolic syndrome | 4 | -0.24 (-0.79, 0.31) | 0%, 0.62 |  |
| CVD | 3 | 0.12 (-0.60, 0.84) | 0%, 0.65 |  |
| **Follow-up duration** |  |  |  | 0.52 |
| ≤12 weeks | 22 | -0.22 (-0.39, -0.06) | 0%, 0.95 |  |
| 12 to ≤24 weeks | 7 | 0.45 (-0.98, 0.07) | 0%, 0.92 |  |
| >24 weeks | 2 | 0.13 (-0.82, 1.07) | 61%, 0.11 |  |
| **Risk of bias** |  |  |  | 0.98 |
| Low | 12 | -0.23 (-0.44, -0.02) | 0%, 0.90 |  |
| Moderate | 9 | -0.21 (-0.50, 0.09) | 0%, 0.77 |  |
| High | 10 | -0.24 (-0.59, 0.10) | 0%, 0.78 |  |
| **Weight status** |  |  |  | 0.49 |
| Normal weight | 25 | -0.15 (-0.41, 0.10) | 0%, 0.93 |  |
| Overweight/obese | 6 | -0.27 (-0.45, -0.08) | 0%, 0.90 |  |
| **Calorie restriction** |  |  |  | 0.83 |
| Yes | 5 | -0.16 (-0.75, 0.42) | 8%, 0.34 |  |
| No | 26 | -0.23 (-0.39, -0.07) | 0%, 0.98 |  |
| **Physical activity** |  |  |  | 0.27 |
| Yes | 3 | 0.18 (-0.67, 1.03) | 0.7%, 0.40 |  |
| No | 28 | -0.24 (-0.40, -0.09) | 0%, 0.98 |  |
| **Behavioral support** |  |  |  | 0.58 |
| Yes | 6 | -0.14 (-0.48, 0.20) | 0%, 0.62 |  |
| No | 25 | -0.25 (-0.42, -0.05) | 0%, 0.98 |  |
| **Funding** |  |  |  | 0.57 |
| Industrial | 19 | -0.20 (-0.37, -0.03) | 0%, 0.86 |  |
| University | 12 | -0.31 (-0.63, 0.01) | 0%, 0.96 |  |
| **Abbreviations:** CVD, cardiovascular disease. |  |  |  |  |

Dose-dependent effect of nuts on DBP are indicated in **Figure 1**, lower panel, and **Table 2**. Levels of DBP decreased linearly and slightly (P_nonlinearity_ = 0.89, P_dose-response_ = 0.02) up to nuts consumption of 80 g/d (MD_80g/d_: -0.80, 95%CI: -1.55, -0.04)

### Grading the evidence

The certainty of evidence was rated using the GRADE approach. For both outcomes, the certainty of evidence was rated moderate for downgrades for serious risk of bias and imprecision, and an upgrade for dose-response gradient (**Supplementary Table 5**).

## Discussion

The present meta-analysis is the first study to investigate the potential dose-dependent effect of nut intake on blood pressure using randomized trials. The analyses indicated that each 20 g/d increment in nuts intake can significantly decrease SBP and DBP by 0.50 and 0.23 mmHg, respectively. The nonlinear dose-response meta-analyses suggested a beneficial dose-dependent effect. Although the certainty of evidence was rated moderate for both outcomes, the effect of nuts on blood pressure levels were small and lower than our MCID (2 mmHg), suggesting the benefit was trivial.

Our findings regarding the effect of nuts intake on blood pressure are in accordance with the results of the previous pairwise meta-analyses. A meta-analysis of 21 randomized trials (11) indicated that nuts consumption can improve SBP in patients with type 2 diabetes by 1.29 mmHg that was comparable to 1.60 mmHg in the present meta-analysis. The study also indicated that pistachios and mixed nuts can improve DBP by 0.80 and 1.19 mmHg, respectively. The greatest DBP reduction in our meta-analysis was about 0.80 mmHg at a dose of 80 g/d. Other pairwise meta-analyses of RCTs have also indicated that dietary interventions to increase the consumption of pistachios (8), cheshaw (9), and almond (10) can improve levels of blood pressure.

However, previous meta-analyses only performed pairwise comparison between intervention and control groups. As mentioned above, there are large variations in the amount of nuts consumed in interventions groups across trials (ranged from 15 to 85 g/d). In addition, the difference between dose of nuts consumption between intervention and control groups within a trial has not been considered in the traditional pairwise meta-analyses. Optimal dose of nuts consumption for reducing blood pressure levels has also not been determined. Dose-response meta-analyses are useful tools for determining the efficacy of increasing dose and selecting optimal dose for intervention (12, 13).

Our analyses indicated that the greatest reduction for SBP was seen at nuts consumption of 40 g/d. An inverse linear association was seen for DBP up to 80 g/d.The favorable effect of nuts on blood pressure levels can be attributed to their high content of mono- and polyunsaturated fatty acids, magnesium, potassium, antioxidants, and dietary fibers (7). Nuts also have anti-inflammatory properties and can improve insulin resistance (58), the two underlying mechanisms for developing hypertension (59, 60). Nuts are rich in arginine and thereby can induce endogenous production of nitric oxide (61), a potent vasodilator (62). Nuts have also been ranked to be the best food group in reducing levels of high density lipoprotein cholesterol (63) and thus, can protect blood vessels against endothelial dysfunction (64).

Despite these favorable effect, our results indicated that the favorable effect of nuts in reducing SBP and DBP was small and was lower than our MCID for blood pressure (2 mmHg). The magnitude of the effect was small in comparison with antihypertensive medications (65), and was modest in comparison with other lifestyle modifications such as aerobic exercise (2.58 to 3.84 mmHg) (66).

In the analysis of SBP, subgroup analyses indicated a stronger effect in patients with type 2 diabetes, as well as in trials that implemented calorie restriction. However, tests for subgroup difference were not significant. In addition, the analyses of DBP did not indicate such stronger effect sizes. There was also no effect modification by physical activity and behavioral support in the analyses of SBP and DBP.

Although nuts have small antihypertensive properties, they are one of the key components of the healthy eating patterns such as the Mediterranean diet and dietary approaches to stop hypertension dietary pattern. The American Heart Association recommends eating four servings (42 grams) of unsalted, unoiled nuts per week for the primary prevention of CVD (67). Epidemiologic research also indicated that each 28 g/d increase in nuts consumption was associated with 7% to 39% lower risk of cardiometabolic disease and all-cause and cause-specific mortality (68). Nuts have a healthy fatty acid profile and are rich in vegetable proteins, fibers, mineral, phytosterols, and carotenoids and thus have anti-inflammatory, antioxidant, and lipid lowering properties (69).

The findings of the present meta-analysis should be interpreted considering the following limitations. Our main limitation is that 26 potentail RCTs could not be included in the present meta-analysis because the dose of nut consumption was not reported in the control groups. This issue can be considered by future trials. Second, of 31 trials included in this review, 22 trials lasted ≤ 12 weeks. More trials are needed to determine whether the favorable effect of nuts on blood pressure levels can exert more than a slight clinical impact in long term. Third, of the trials, only 12 trials have been rated to have low risk of bias. However, subgroup analyses in studies with low risk of bias confirmed the main findings. Fifth, the compliance of participants to study interventions was rarely reported. Thus, we were unable to assess attrition bias in the included trials and its potential impact on the overall results. Sixth, due to lack of trials conducted exclusively in patients with hypertension, we were unable to assess the effects in hypertensive individuals.

## Conclusions

The present dose-response meta-analysis of randomized trials gave a good indication that nut consumption can result in small improvement in blood pressure levels in adults. The results suggested a dose-dependent effect, with the greatest reduction being observed at nuts consumption of 40 g/d for SBP and 80 g/d for DBP. Well-designed trials are needed to confirm the findings in long term follow-up.

## Data Availability

The data used in the present study will be provided upon request.

## References

1. Murray CJ, Aravkin AY, Zheng P, Abbafati C, Abbas KM, Abbasi-Kangevari M, Abd-Allah F, Abdelalim A, Abdollahi M, Abdollahpour I. Global burden of 87 risk factors in 204 countries and territories, 1990–2019: a systematic analysis for the Global Burden of Disease Study 2019. The Lancet 2020;396(10258):1223–49.

2. Hajjar I, Kotchen TA. Trends in prevalence, awareness, treatment, and control of hypertension in the United States, 1988-2000. Jama 2003;290(2):199–206.

3. Forouzanfar MH, Liu P, Roth GA, Ng M, Biryukov S, Marczak L, Alexander L, Estep K, Abate KH, Akinyemiju TF. Global burden of hypertension and systolic blood pressure of at least 110 to 115 mm Hg, 1990-2015. Jama 2017;317(2):165–82.

4. Mills KT, Stefanescu A, He J. The global epidemiology of hypertension. Nature Reviews Nephrology 2020;16(4):223–37.

5. Jayedi A, Rashidy-Pour A, Khorshidi M, Shab-Bidar S. Body mass index, abdominal adiposity, weight gain and risk of developing hypertension: a systematic review and dose–response meta-analysis of more than 2.3 million participants. Obesity reviews 2018;19(5):654–67.

6. Schwingshackl L, Schwedhelm C, Hoffmann G, Knüppel S, Iqbal K, Andriolo V, Bechthold A, Schlesinger S, Boeing H. Food groups and risk of hypertension: a systematic review and dose-response meta-analysis of prospective studies. Advances in nutrition 2017;8(6):793–803.

7. Casas-Agustench P, Lopez-Uriarte P, Ros E, Bullo M, Salas-Salvado J. Nuts, hypertension and endothelial function. Nutrition, Metabolism and Cardiovascular Diseases 2011;21:S21–S33.

8. Asbaghi O, Hadi A, Campbell MS, Venkatakrishnan K, Ghaedi E. Effects of pistachios on anthropometric indices, inflammatory markers, endothelial function, and blood pressure in adults: A systematic review and meta-analysis of randomized controlled trials. British Journal of Nutrition 2020:1–27.

9. Jalali M, Karamizadeh M, Ferns GA, Zare M, Moosavian SP, Akbarzadeh M. The effects of cashew nut intake on lipid profile and blood pressure: a systematic review and meta-analysis of randomized controlled trials. Complementary Therapies in Medicine 2020;50:102387.

10. Li Z, Bhagavathula AS, Batavia M, Clark C, Abdulazeem HM, Rahmani J, Yin F. The effect of almonds consumption on blood pressure: A systematic review and dose-response meta-analysis of randomized control trials. Journal of King Saud University-Science 2020;32(2):1757–63.

11. Mohammadifard N, Salehi-Abargouei A, Salas-Salvadó J, Guasch-Ferré M, Humphries K, Sarrafzadegan N. The effect of tree nut, peanut, and soy nut consumption on blood pressure: a systematic review and meta-analysis of randomized controlled clinical trials. The American journal of clinical nutrition 2015;101(5):966–82.

12. Bretz F, Hsu J, Pinheiro J, Liu Y. Dose finding–a challenge in statistics. Biometrical Journal: Journal of Mathematical Methods in Biosciences 2008;50(4):480–504.

13. Bretz F, Pinheiro JC, Branson M. Combining multiple comparisons and modeling techniques in dose-response studies. Biometrics 2005;61(3):738–48.

14. Crippa A, Orsini N. Dose-response meta-analysis of differences in means. BMC medical research methodology 2016;16(1):1–10.

15. Page MJ, Moher D, Bossuyt PM, Boutron I, Hoffmann TC, Mulrow CD, Shamseer L, Tetzlaff JM, Akl EA, Brennan SE. PRISMA 2020 explanation and elaboration: updated guidance and exemplars for reporting systematic reviews. bmj 2021;372.

16. Liu K, Hui S, Wang B, Kaliannan K, Guo X, Liang L. Comparative effects of different types of tree nut consumption on blood lipids: a network meta-analysis of clinical trials. The American journal of clinical nutrition 2020;111(1):219–27.

17. Higgins JP, Altman DG, Gøtzsche PC, Jüni P, Moher D, Oxman AD, Savović J, Schulz KF, Weeks L, Sterne JA. The Cochrane Collaboration’s tool for assessing risk of bias in randomised trials. Bmj 2011;343.

18. Chandler J, Cumpston M, Li T, Page M, Welch V. Cochrane handbook for systematic reviews of interventions. Hoboken: Wiley 2019.

19. Higgins JP, Deeks JJ. Selecting studies and collecting data. Cochrane handbook for systematic reviews of interventions: Cochrane book series 2008:151–85.

20. Furukawa TA, Barbui C, Cipriani A, Brambilla P, Watanabe N. Imputing missing standard deviations in meta-analyses can provide accurate results. Journal of clinical epidemiology 2006;59(1):7–10.

21. DerSimonian R, Laird N. Meta-analysis in clinical trials. Controlled clinical trials 1986;7(3):177–88.

22. Egger M, Smith GD, Schneider M, Minder C. Bias in meta-analysis detected by a simple, graphical test. Bmj 1997;315(7109):629–34.

23. Begg CB, Mazumdar M. Operating characteristics of a rank correlation test for publication bias. Biometrics 1994:1088–101.

24. Higgins JP, Savović J, Page MJ, Elbers RG, Sterne JA. Assessing risk of bias in a randomized trial. Cochrane handbook for systematic reviews of interventions 2019:205–28.

25. Guyatt G, Oxman AD, Akl EA, Kunz R, Vist G, Brozek J, Norris S, Falck-Ytter Y, Glasziou P, Debeer H. GRADE guidelines: 1. Introduction—GRADE evidence profiles and summary of findings tables. Journal of clinical epidemiology 2011;64(4):383–94.

26. Ge L, Sadeghirad B, Ball GD, da Costa BR, Hitchcock CL, Svendrovski A, Kiflen R, Quadri K, Kwon HY, Karamouzian M. Comparison of dietary macronutrient patterns of 14 popular named dietary programmes for weight and cardiovascular risk factor reduction in adults: systematic review and network meta-analysis of randomised trials. bmj 2020;369.

27. Al Abdrabalnabi A, Rajaram S, Bitok E, Oda K, Beeson WL, Kaur A, Cofán M, Serra-Mir M, Roth I, Ros E. Effects of supplementing the usual diet with a daily dose of walnuts for two years on metabolic syndrome and its components in an elderly cohort. Nutrients 2020;12(2):451.

28. Campos V, Portal V, Markoski M, Quadros A, Bersch-Ferreira Â, Garavaglia J, Marcadenti A. Effects of a healthy diet enriched or not with pecan nuts or extra-virgin olive oil on the lipid profile of patients with stable coronary artery disease: a randomised clinical trial. Journal of Human Nutrition and Dietetics 2020;33(3):439–50.

29. Casas-Agustench P, López-Uriarte P, Bulló M, Ros E, Cabré-Vila J, Salas-Salvadó J. Effects of one serving of mixed nuts on serum lipids, insulin resistance and inflammatory markers in patients with the metabolic syndrome. Nutrition, metabolism and cardiovascular diseases 2011;21(2):126–35.

30. Chen C-M, Liu J-F, Li S-C, Huang C-L, Hsirh A-T, Weng S-F, Chang M-L, Li H-T, Mohn E, Chen CO. Almonds ameliorate glycemic control in Chinese patients with better controlled type 2 diabetes: a randomized, crossover, controlled feeding trial. Nutrition & metabolism 2017;14(1):1–12.

31. Chen CO, Holbrook M, Duess M-A, Dohadwala MM, Hamburg NM, Asztalos BF, Milbury PE, Blumberg JB, Vita JA. Effect of almond consumption on vascular function in patients with coronary artery disease: a randomized, controlled, cross-over trial. Nutrition journal 2015;14(1):1–11.

32. de Souza RGM, Gomes AC, de Castro IA, Mota JF. A baru almond-enriched diet reduces abdominal adiposity and improves high-density lipoprotein concentrations: a randomized, placebo-controlled trial. Nutrition 2018;55-56:154–60.

33. Dhillon J, Tan S-Y, Mattes RD. Almond consumption during energy restriction lowers truncal fat and blood pressure in compliant overweight or obese adults. The Journal of nutrition 2016;146(12):2513–9.

34. Din JN, Aftab SM, Jubb AW, Carnegy FH, Lyall K, Sarma J, Newby DE, Flapan AD. Effect of moderate walnut consumption on lipid profile, arterial stiffness and platelet activation in humans. European journal of clinical nutrition 2011;65(2):234–9.

35. Dusanov S, Svendsen M, Ruzzin J, Kiviranta H, Gulseth HL, Klemsdal TO, Tonstad S. Effect of fatty fish or nut consumption on concentrations of persistent organic pollutants in overweight or obese men and women: A randomized controlled clinical trial. Nutrition, Metabolism and Cardiovascular Diseases 2020;30(3):448–58.

36. Foster GD, Shantz KL, Vander Veur SS, Oliver TL, Lent MR, Virus A, Szapary PO, Rader DJ, Zemel BS, Gilden-Tsai A. A randomized trial of the effects of an almond-enriched, hypocaloric diet in the treatment of obesity. The American journal of clinical nutrition 2012;96(2):249–54.

37. Ghadimi Nouran M, Kimiagar M, Abadi A, Mirzazadeh M, Harrison G. Peanut consumption and cardiovascular risk. Public Health Nutr 2010;13(10):1581–6.

38. Hernández-Alonso P, Salas-Salvadó J, Baldrich-Mora M, Juanola-Falgarona M, Bulló M. Beneficial effect of pistachio consumption on glucose metabolism, insulin resistance, inflammation, and related metabolic risk markers: a randomized clinical trial. Diabetes care 2014;37(11):3098–105.

39. Jamshed H, Sultan FAT, Iqbal R, Gilani AH. Dietary almonds increase serum HDL cholesterol in coronary artery disease patients in a randomized controlled trial. The Journal of nutrition 2015;145(10):2287–92.

40. Katz DL, Davidhi A, Ma Y, Kavak Y, Bifulco L, Njike VY. Effects of walnuts on endothelial function in overweight adults with visceral obesity: a randomized, controlled, crossover trial. Journal of the American College of Nutrition 2012;31(6):415–23.

41. Ma Y, Njike VY, Millet J, Dutta S, Doughty K, Treu JA, Katz DL. Effects of walnut consumption on endothelial function in type 2 diabetic subjects: a randomized controlled crossover trial. Diabetes care 2010;33(2):227–32.

42. McKay DL, Eliasziw M, Chen C, Blumberg JB. A pecan-rich diet improves cardiometabolic risk factors in overweight and obese adults: a randomized controlled trial. Nutrients 2018;10(3):339.

43. Mohan V, Gayathri R, Jaacks LM, Lakshmipriya N, Anjana RM, Spiegelman D, Jeevan RG, Balasubramaniam KK, Shobana S, Jayanthan M. Cashew nut consumption increases HDL cholesterol and reduces systolic blood pressure in Asian Indians with type 2 diabetes: a 12-week randomized controlled trial. The Journal of nutrition 2018;148(1):63–9.

44. Mukuddem-Petersen J, Stonehouse Oosthuizen W, Jerling JC, Hanekom SM, White Z. Effects of a high walnut and high cashew nut diet on selected markers of the metabolic syndrome: a controlled feeding trial. Br J Nutr 2007;97(6):1144–53.

45. Ndanuko RN, Tapsell LC, Charlton KE, Neale EP, Batterham MJ. Effect of individualised dietary advice for weight loss supplemented with walnuts on blood pressure: the HealthTrack study. European Journal of Clinical Nutrition 2018;72(6):894–903.

46. Njike VY, Ayettey R, Petraro P, Treu JA, Katz DL. Walnut ingestion in adults at risk for diabetes: effects on body composition, diet quality, and cardiac risk measures. BMJ Open Diabetes Res Care 2015;3(1):e000115.

47. Olmedilla-Alonso B, Granado-Lorencio F, Herrero-Barbudo C, Blanco-Navarro I, Blázquez-García S, Pérez-Sacristán B. Consumption of restructured meat products with added walnuts has a cholesterol-lowering effect in subjects at high cardiovascular risk: a randomised, crossover, placebo-controlled study. J Am Coll Nutr 2008;27(2):342–8.

48. Parham M, Heidari S, Khorramirad A, Hozoori M, Hosseinzadeh F, Bakhtyari L, Vafaeimanesh J. Effects of pistachio nut supplementation on blood glucose in patients with type 2 diabetes: a randomized crossover trial. Rev Diabet Stud 2014;11(2):190–6.

49. Rock CL, Flatt SW, Barkai H-S, Pakiz B, Heath DD. Walnut consumption in a weight reduction intervention: effects on body weight, biological measures, blood pressure and satiety. Nutrition Journal 2017;16(1):76.

50. Rock CL, Zunshine E, Nguyen HT, Perez AO, Zoumas C, Pakiz B, White MM. Effects of Pistachio Consumption in a Behavioral Weight Loss Intervention on Weight Change, Cardiometabolic Factors, and Dietary Intake. Nutrients 2020;12(7).

51. Spaccarotella KJ, Kris-Etherton PM, Stone WL, Bagshaw DM, Fishell VK, West SG, Lawrence FR, Hartman TJ. The effect of walnut intake on factors related to prostate and vascular health in older men. Nutrition journal 2008;7:13-.

52. Tey SL, Gray AR, Chisholm AW, Delahunty CM, Brown RC. The dose of hazelnuts influences acceptance and diet quality but not inflammatory markers and body composition in overweight and obese individuals. J Nutr 2013;143(8):1254–62.

53. Tindall AM, Petersen KS, Skulas-Ray AC, Richter CK, Proctor DN, Kris-Etherton PM. Replacing Saturated Fat With Walnuts or Vegetable Oils Improves Central Blood Pressure and Serum Lipids in Adults at Risk for Cardiovascular Disease: A Randomized Controlled-Feeding Trial. J Am Heart Assoc 2019;8(9):e011512.

54. Wang X, Li Z, Liu Y, Lv X, Yang W. Effects of pistachios on body weight in Chinese subjects with metabolic syndrome. Nutrition journal 2012;11:20-.

55. West SG, Gebauer SK, Kay CD, Bagshaw DM, Savastano DM, Diefenbach C, Kris-Etherton PM. Diets containing pistachios reduce systolic blood pressure and peripheral vascular responses to stress in adults with dyslipidemia. Hypertension 2012;60(1):58–63.

56. Wien MA, Sabaté JM, Iklé DN, Cole SE, Kandeel FR. Almonds vs complex carbohydrates in a weight reduction program. Int J Obes Relat Metab Disord 2003;27(11):1365–72.

57. Wu H, Pan A, Yu Z, Qi Q, Lu L, Zhang G, Yu D, Zong G, Zhou Y, Chen X, et al. Lifestyle counseling and supplementation with flaxseed or walnuts influence the management of metabolic syndrome. The Journal of nutrition 2010;140(11):1937–42.

58. Casas-Agustench P, Bulló M, Salas-Salvadó J. Nuts, inflammation and insulin resistance. Asia Pacific journal of clinical nutrition 2010;19(1):124.

59. Ferrannini E, Buzzigoli G, Bonadonna R, Giorico MA, Oleggini M, Graziadei L, Pedrinelli R, Brandi L, Bevilacqua S. Insulin resistance in essential hypertension. New England Journal of Medicine 1987;317(6):350–7.

60. Jayedi A, Rahimi K, Bautista LE, Nazarzadeh M, Zargar MS, Shab-Bidar S. Inflammation markers and risk of developing hypertension: a meta-analysis of cohort studies. Heart 2019;105(9):686–92.

61. Ros E, Tapsell LC, Sabaté J. Nuts and berries for heart health. Current atherosclerosis reports 2010;12(6):397–406.

62. Bode-Böger SM, Böger RH, Alfke H, Heinzel D, Tsikas D, Creutzig A, Alexander K, Frölich JrC. L-Arginine induces nitric oxide–dependent vasodilation in patients with critical limb ischemia: a randomized, controlled study. Circulation 1996;93(1):85–90.

63. Schwingshackl L, Hoffmann G, Iqbal K, Schwedhelm C, Boeing H. Food groups and intermediate disease markers: a systematic review and network meta-analysis of randomized trials. The American journal of clinical nutrition 2018;108(3):576–86.

64. Gradinaru D, Borsa C, Ionescu C, Prada GI. Oxidized LDL and NO synthesis—biomarkers of endothelial dysfunction and ageing. Mechanisms of ageing and development 2015;151:101–13.

65. Patel HC, Hayward C, Ozdemir BA, Rosen SD, Krum H, Lyon AR, Francis DP, Di Mario C. Magnitude of blood pressure reduction in the placebo arms of modern hypertension trials: implications for trials of renal denervation. Hypertension 2015;65(2):401–6.

66. Whelton SP, Chin A, Xin X, He J. Effect of aerobic exercise on blood pressure: a meta-analysis of randomized, controlled trials. Annals of internal medicine 2002;136(7):493–503.

67. Arnett DK, Blumenthal RS, Albert MA, Buroker AB, Goldberger ZD, Hahn EJ, Himmelfarb CD, Khera A, Lloyd-Jones D, McEvoy JW. 2019 ACC/AHA guideline on the primary prevention of cardiovascular disease: executive summary: a report of the American College of Cardiology/American Heart Association Task Force on Clinical Practice Guidelines. Journal of the American College of Cardiology 2019;74(10):1376–414.

68. Aune D, Keum N, Giovannucci E, Fadnes LT, Boffetta P, Greenwood DC, Tonstad S, Vatten LJ, Riboli E, Norat T. Nut consumption and risk of cardiovascular disease, total cancer, all-cause and cause-specific mortality: a systematic review and dose-response meta-analysis of prospective studies. BMC medicine 2016;14(1):1–14.

69. De Souza RGM, Schincaglia RM, Pimentel GD, Mota JF. Nuts and human health outcomes: A systematic review. Nutrients 2017;9(12):1311.

