## Supplementary Tables 1-5 and Supplementary Figures 1-5 for "Dose-dependent effect of nuts on blood pressure: a systematic review and meta-analysis of randomized controlled trials"

**Supplementary Table** 1. Search strategy to find potential eligible trials for inclusion in dose-response meta-analysis of nuts and blood pressure.

**PubMed**: March 04, 2021 (213)

(Nuts [Mesh] OR nut* [Title/Abstract] OR almond* [Title/Abstract] OR walnut* [Title/Abstract] OR "mixed nuts" [Title/Abstract] OR hazelnut* [Title/Abstract] OR pistachio* [Title/Abstract] OR peanut* [Title/Abstract] OR cashew* [Title/Abstract] OR macadamia [Title/Abstract] OR "Brazil nuts" [Title/Abstract] OR "peanut butter" [Title/Abstract]) **AND** ("Random Allocation"[Mesh] OR "Single-Blind Method"[Mesh] OR "Double-Blind Method"[Mesh] OR "Cross-Over Studies"[Mesh] OR "Clinical Trials as Topic"[Mesh] OR RCT[Title/Abstract] OR "Intervention Studies"[Title/Abstract] OR intervention[Title/Abstract] OR "controlled trial"[Title/Abstract] OR randomized[Title/Abstract] OR randomised[Title/Abstract] OR random[Title/Abstract] OR randomly[Title/Abstract] OR placebo[Title/Abstract] OR assignment[Title/Abstract]) **AND** “Blood pressure” [Mesh] OR Hypertension [Mesh] OR "blood pressure" [Title/Abstract] OR "systolic blood pressure" [Title/Abstract] OR "diastolic blood pressure" [Title/Abstract] OR sbp [Title/Abstract] OR dbp [Title/Abstract] OR hypertension [Title/Abstract] OR hypertens* [Title/Abstract] OR "Systolic Pressure" [Title/Abstract] OR "Diastolic Pressure" [Title/Abstract] OR "High Blood Pressure" [Title/Abstract] OR "Arterial Pressure" [Title/Abstract] OR "Arterial Blood Pressure" [Title/Abstract])

**Scopus**: March 04, 2021 (3602)

**ISI Web of Siences**: March 04, 2021 (237)

**Total**: 4502

**Supplementary Table 2**. List of studies excluded via full text assessment.

1. Dose in the control group not reported (n=26) (1-26)

2. Not relevant intervention (n=26) (27-52)

3. Not relevant outcome (n=10) (53-61)

4. Duplicate (n=8) (62-69)

5. Active control (n=3) (70-72)

6. Quasi experimental (n=2) (73, 74)

7. Not sufficient information (n=2) (75, 76)

**Supplementary Table 2**. Characteristics of trials included in dose-response meta-analysis of nuts consumption and blood pressure.

| Reference, country | Participants  (n) | Age range | Study design (duration, wk) | Treatment  (dose) | Control | BMI range (mean, kg/m^2^) |
| --- | --- | --- | --- | --- | --- | --- |
| Al Abdrabalnabi, 2020,  US | Healthy elderly individuals  (636) | 62-79 | Parallel  (108) | Walnut  (56 g/d) | Usual nut-free diet | 27.8 |
| Campos, 2019  Brazil | Patients with stable coronary artery disease  (n=135) | 40-80 | Parallel  (12) | Healthy diet + Pecan  (30 g/d) | Healthy nut-free diet according to the nutritional guidelines | 29.3 |
| Casas-Agustench, 2009  Spain | Patients with the metabolic syndrome (n=50) | 18-65 | Parallel  (12) | Mixed nuts  (30 g/d) | Healthy nut-free diet | 30.8 |
| Chen, 2015  US | Patients with coronary artery disease  (n=45) | 21-80 | Cross-over (12) | National Cholesterol Education Program (NCEP) + Almond  (85 g/d) | NCEP Step 1 nut-free diet | 20-41  (30.2) |
| Chen, 2017  Taiwan | Patients with type 2 diabetes  (n=33) | 40-70 | Cross-over (12) | NCEP step II + Almond  (60 g/d) | NCEP step II nut-free diet | 24-35  (25.5) |
| Dhillon, 2016  US | Overweight or obese adults  (n=66) | 18-60 | Parallel  (12) | Almond (60 g/d) | A nut-free diet | 25-40 |
| Din, 2011  UK | Healthy males  (n=30) | 23 ± 3 | Cross-over  (4) | Walnut  (15 g/d) | A nut-free diet | 24.5 |
| Dusanov, 2019  Norway | Overweight or obese men and women  (n=23) | 35-70 | Parallel  (24) | Mixed nuts  (30 g/d) | Usual nut-free diet | 25-38 |
| Foster, 2012  US | Overweight and obese individuals  (n=123) | 18-75 | Parallel  (72) | A hypocaloric diet + Almond  (56 g/d) | A hypocaloric nut-free diet | 34 |
| Hern´andez-Alonso, 2014  Spain | Prediabetic subjects  (n=54) | 25-65 | Cross-over  (12) | Pistachio  (57 g/d) | Usual nut-free diet | 28.9 |
| Jamshed, 2015  Pakistan | Patients with coronary artery disease  (n=75) | 32-86 | Parallel  (16) | Almond  (10 g/d) | Usual nut-free diet | 27.3 |
| Katz, 2013  US | Overweight adults with elevated waist circumference and 1 or more additional signs of metabolic syndrome  (n=40) | 30-75 | Cross-over  (12) | Walnut  (56 g/d) | *ad libitum* nut-free diet | >25 |
| Ma, 2010  US | Participants with type 2 diabetes  (n=21) | 30-75 | Cross-over  (8) | Walnut  (56 g/d) | *ad libitum* nut-free diet | 32.5 |
| Mckay, 2018  US | Metabolically at-risk, nonsmoking men and postmenopausal women  (n=26) | >45 | Cross-over  (4) | Pecan nut  (47.5 g/d) | Isocaloric nut-free control diet | 25-35 |
| Mohan, 2018  India | Patients with type 2 diabetes  (n=269) | 30-65 | Parallel  (12) | Cashew  (30 g/d) | A standard diabetic nut free diet | 26 |
| Mukuddem-Petersen, 2007  South Africa | Subjects having the metabolic syndrome  (n=63) | 45 | Parallel  (8) | Cashew  (85.5 g/d)  Walnut  (85.5 g/d) | Nut free diet | 35.2 |
| Ndanuko, 2018  Australia | Apparently healthy adults  (n=144) | 25-54 | Parallel  (12) | Walnut  (30 g/d) | General advice and nut-free diet | 20-40 |
| Njike, 2015  US | Adults at risk for diabetes  (n=52) | 25-75 | Parallel  (24) | Walnut  (56 g/d) | Nut-free diet | ≥25 |
| Nouran, 2009  Iran | Hypercholesterolemic men  (n=44) | 25-65 | Parallel  (24) | Peanut  (77 g/d) | Nut-free diet | 27.5 |
| Olmedilla-Alonso, 2014  Spain | Subjects at high cardiovascular risk  (n=25) | 45-65 | Parallel  (4) | Walnut  (19 g/d) | Nut-free diet | >25 |
| Parham, 2014  Iran | Patients with type 2 diabetes  (n=44) | 51.5 | Cross-over  (24) | Pistachio  (50 g/d) | Nut-free diet | 32.2 |
| Rock, 2017  US | Overweight and obese men and women  (n=100) | >21 | Parallel  (24) | Standard reduced energy-density diet + Walnut  (35 g/d) | Standard reduced energy-  density nut-free diet | 27-40 |
| Rock, 2020  US | Non-diabetic overweight/obese adults  (n=104) | >21 | Parallel  (16) | Pistachio  (42 g/d) | Usual nut-free diet | 27-40 |
| Souza, 2018  Brazil | Overweight and obese women  (n=46) | 20-59 | Parallel  (8) | Almond  (20 g/d) | Nut-free diet | 25-40 |
| Spaccarotella, 2008  US | Men at risk for prostate cancer  (n=42) | 45-75 | Parallel  (8) | Walnut  (75 g/d) | Nut-free diet | 29.4 |
| Tey, 2013  New Zealand | Overweight and obese individuals  (n=107) | 18-65 | Parallel  (12) | Hazelnut  (30 and 60 gr/d) | Nut-free diet | ≥25 |
| Tindall, 2019  US | Adults at risk for cardiovascular disease (n=45) |  | Cross-over  (12) | Walnut  (78 g/d) | Nut-free diet | 25-40 |
| Wang, 2019  China | Subjects with metabolic syndrome  (n=60) | 25-65 | Parallel  (6) | Pistachio  (42 and 70 gr/d) | Nut-free diet | 28 |
| West, 2012  US | Adults with dyslipidemia  (n=28) | 50 | Cross-over  (4) | Pistachio  (35 and 69 gr/d) | Nut-free diet | 21-35 |
| Wien, 2003  US | Overweight and obese adults  (n=65) | 27-79 | Parallel  (16) | Almond  (70 g/d) | Nut-free diet | 27-55 |
| Wu, 2010  China | Adults with metabolic syndrome  (n=189) | 48.4 | Parallel  (12) | Walnut  (30 g/d) | Nut-free diet | 25.4 |

**Supplementary Table 4.** Quality of trials included in the meta-analysis of the effects of nuts on blood pressure.

| Study, year | Random Sequence Generation | Allocation concealment | Blinding of participants and personnel | Blinding of outcome assessment | Incomplete outcome data | Selective outcome reporting | Other sources of bias | Overall quality |
| --- | --- | --- | --- | --- | --- | --- | --- | --- |
| Abdrabalnabi, 2019 | L | U | L | L | L | L | L | Good |
| Campos, 2019 | L | L | L | U | L | L | L | Good |
| Casas-Agustench, 2011 | L | L | U | U | L | L | L | Fair |
| Chen, 2015 | L | U | U | L | L | L | L | Fair |
| Chen, 2017 | L | U | U | L | L | L | L | Fair |
| Dhillon, 2016 | L | H | H | L | L | L | L | Poor |
| Din, 2010 | L | L | L | L | L | L | L | Good |
| Dusanov, 2019 | L | U | U | L | L | L | L | Fair |
| Foster, 2012 | L | U | H | U | L | L | L | Poor |
| Hernandez-Alonso, 2014 | L | U | H | H | L | L | L | Poor |
| Jamshed, 2015 | L | U | H | L | L | L | L | Fair |
| Katz, 2013 | L | U | L | L | L | L | L | Good |
| Ma, 2010 | L | U | L | U | L | L | L | Good |
| McKay, 2018 | L | L | L | U | U | L | L | Fair |
| Mohan, 2018 | L | L | H | U | L | L | L | Fair |
| Mukuddem-Petersen, 2007 | L | L | U | U | L | L | L | Fair |
| Ndanuko, 2018 | L | L | L | L | L | L | L | Good |
| Njike, 2017 | L | L | L | L | L | L | L | Good |
| Nouran, 2009 | U | U | H | U | L | L | L | Poor |
| Olmedilla-Alonso, 2013 | L | U | L | L | U | L | L | Fair |
| Parham, 2014 | L | U | H | H | L | L | L | Poor |
| Rock, 2017 | L | U | L | L | L | L | L | Good |
| Rock, 2020 | L | U | L | L | L | L | L | Good |
| Souza, 2018 | L | L | U | L | L | L | L | Good |
| Spaccarotella, 2008 | L | U | H | U | L | H | U | Poor |
| Tey, 2015 | L | U | U | L | L | L | U | Poor |
| Tindall, 2019 | L | L | L | L | L | L | L | Good |
| Wang, 2012 | U | U | U | H | U | U | U | Poor |
| West, 2012 | U | U | H | U | U | L | L | Poor |
| Wien, 2013 | L | L | L | U | L | L | L | Good |
| Wu 2010 | L | U | H | L | L | L | U | Poor |

**Supplementary Table 5**. GRADE evidence table for the effects of nuts on blood pressure.

| **Certainty assessment** | | | | | | | **№ of patients** | | **Mean difference (95%CI)** | **Certainty** | **Importance** |
| --- | --- | --- | --- | --- | --- | --- | --- | --- | --- | --- | --- |
| **№ of studies** | **Study design** | **Risk of bias** | **Inconsistency** | **Indirectness** | **Imprecision** | **Other considerations** | **[intervention]** | **[comparison]** |  |  |  |
| **Systolic Blood Pressure** | | | | | | | | | | | |
| 31 | randomised trials | serious ^a^ | not serious | not serious | serious ^b^ | dose response gradient | 1380 | 1404 | MD **0.5 mmHg lower** (0.79 lower to 0.29 lower) | ⨁⨁⨁◯ MODERATE | IMPORTANT |
| **Diastolic Blood Pressure** | | | | | | | | | | | |
| 31 | randomised trials | serious ^c^ | not serious | not serious | serious ^d^ | dose response gradient | 1380 | 1404 | **0.23 mmHg lower** (0.38 lower to 0.08 lower) | ⨁⨁⨁◯ MODERATE | IMPORTANT |

**CI:** Confidence interval; **MD:** Mean difference

##### Explanations

a. Serious risk of bias since only 12 trials out of 31 trials were rated to have low risk of bias. Downgraded. / b. Serious imprecision since point estimate did not surpass our MCID for SBP (2 mmHg). Downgraded. / c. Serious risk of bias since only 12 trials out of 31 trials were rated to have low risk of bias. Downgraded. / d. Serious imprecision since point estimate did not surpass our MCID for DBP (2 mmHg). Downgraded.

Studies included in quantitative synthesis (meta-analysis)
(n =31)

Studies included in qualitative synthesis
(n =31)

Full-text articles assessed for eligibility
(n = 108)

Records excluded
(n = 4103)

Records screened
(n = 4211)

Records after duplicates removed
(n = 4211)

### Identification

### Eligibility

### Included

### Screening

Records identified through database searching
(n = 4502)

Additional records identified through other sources
(n = 0)

Full-text articles excluded, with reasons
(n = 77): No dose in control group (n=26)	Not relevant intervention (n=26)	Not relevant outcome (n=10)	Duplicates (n=8)	Active control (n=3)	Quasi experimental (n=2)	Not sufficient information (n=2)

**Supplemental Figure 1**. Literature search and study selection process.

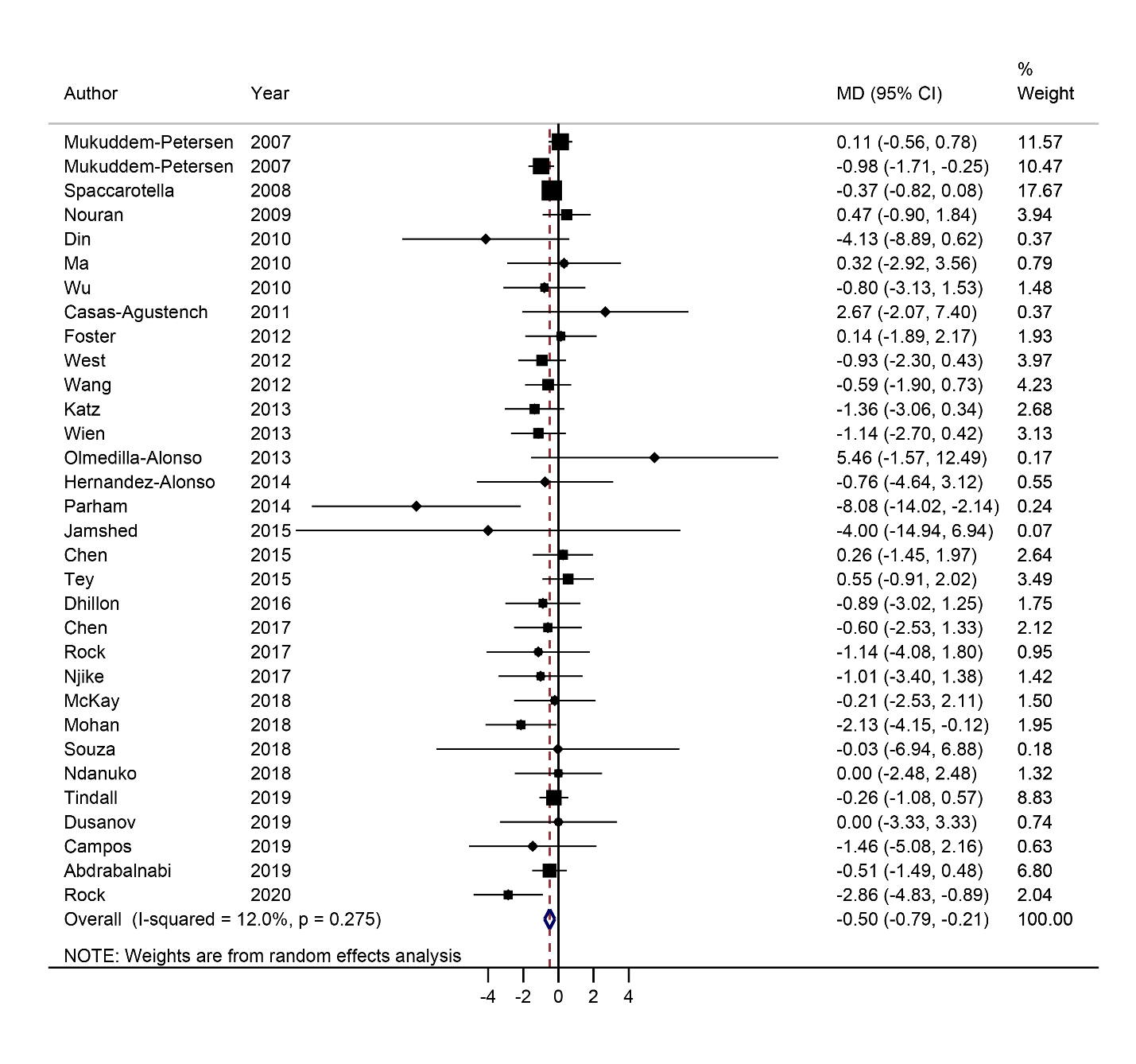

**Supplementary Figure 2**. Mean difference and 95%CI of change in systolic blood pressure for each 20 g/d increment in nuts consumption. MD, mean difference.

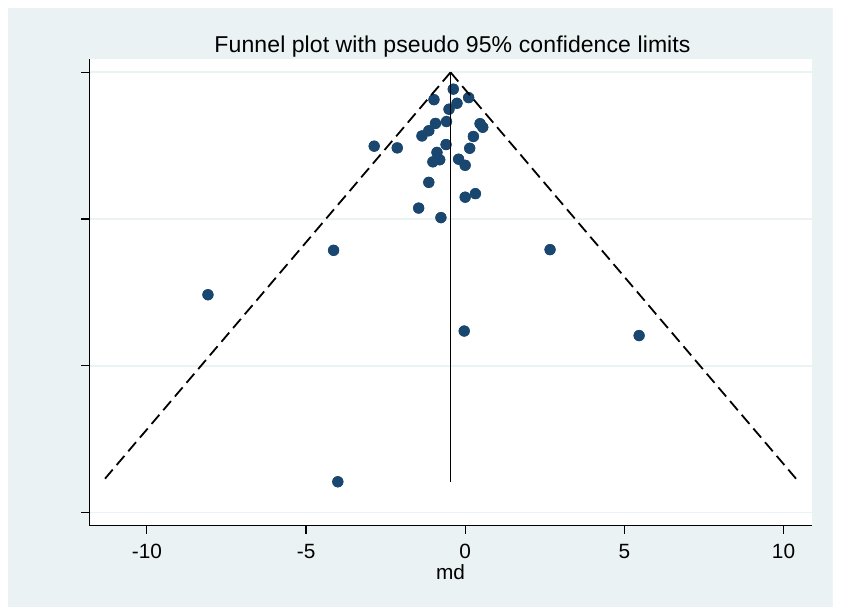

**Supplementary Figure 3**. Funnel plot of the effects of nuts on systolic blood pressure. se, standard error.

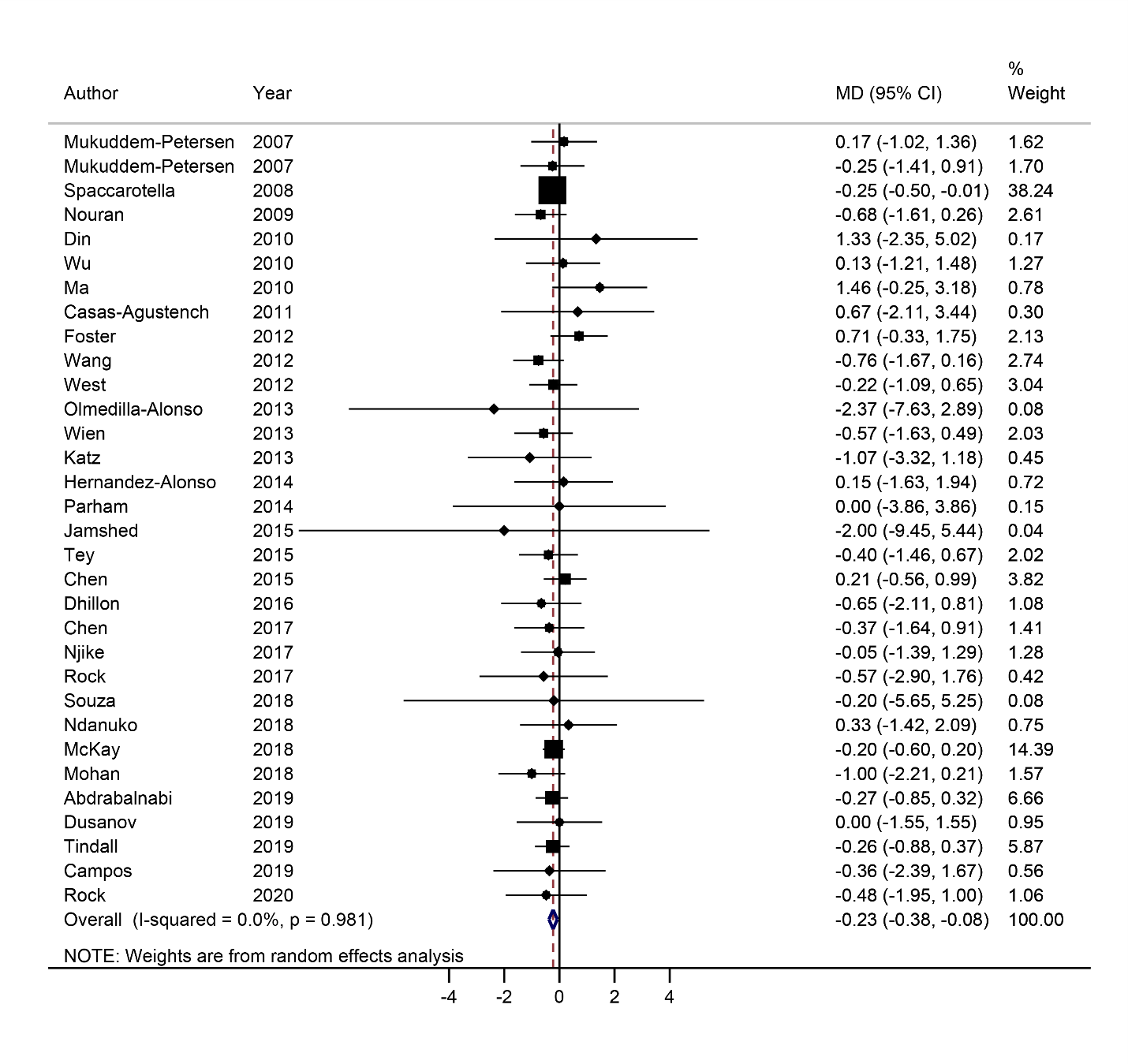

**Supplementary Figure 4**. Mean difference and 95%CI of change in diastolic blood pressure for each 20 g/d increment in nuts consumption. MD, mean difference.

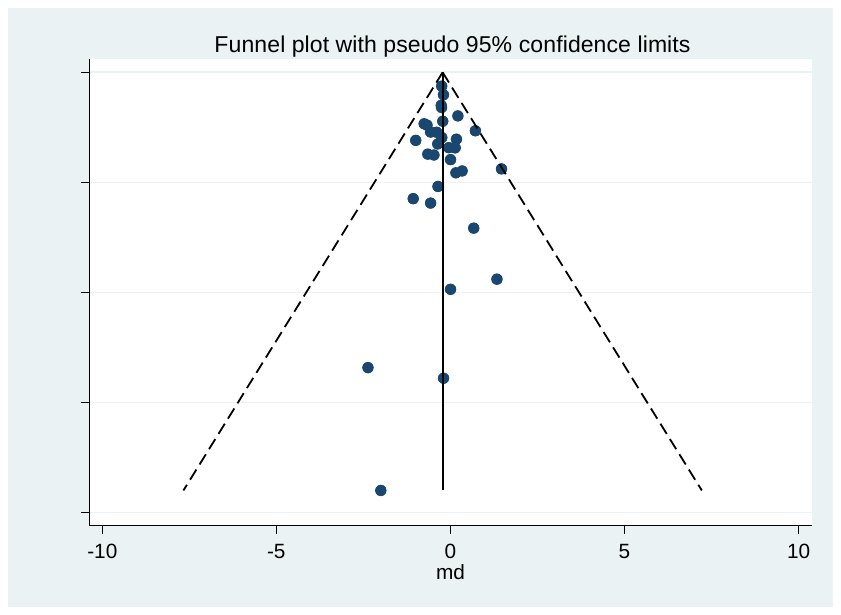

**Supplementary Figure 5**. Funnel plot of the effects of nuts on diastolic blood pressure. se, standard error.

**References**:

1.	Abazarfard Z, Salehi M, Keshavarzi S. The effect of almonds on anthropometric measurements and lipid profile in overweight and obese females in a weight reduction program: A randomized controlled clinical trial. Journal of research in medical sciences : the official journal of Isfahan University of Medical Sciences 2014;19(5):457-64.

2.	Abbaspour N, Roberts T, Hooshmand S, Kern M, Hong MY. Mixed Nut Consumption May Improve Cardiovascular Disease Risk Factors in Overweight and Obese Adults. Nutrients 2019;11(7). doi: 10.3390/nu11071488.

3.	Carvalho RF, Huguenin GV, Luiz RR, Moreira AS, Oliveira GM, Rosa G. Intake of partially defatted Brazil nut flour reduces serum cholesterol in hypercholesterolemic patients--a randomized controlled trial. Nutr J 2015;14:59. doi: 10.1186/s12937-015-0036-x.

4.	Chisholm A, Mc Auley K, Mann J, Williams S, Skeaff M. Cholesterol lowering effects of nuts compared with a Canola oil enriched cereal of similar fat composition. Nutrition, metabolism, and cardiovascular diseases : NMCD 2005;15(4):284-92. doi: 10.1016/j.numecd.2005.01.006.

5.	Coates AM, Morgillo S, Yandell C, Scholey A. Effect of a 12-Week Almond-Enriched Diet on Biomarkers of Cognitive Performance, Mood, and Cardiometabolic Health in Older Overweight Adults. 2020;12(4). doi: 10.3390/nu12041180.

6.	Dikariyanto V, Smith L, Francis L, Robertson M, Kusaslan E, O'Callaghan-Latham M, Palanche C, D'Annibale M, Christodoulou D, Basty N, et al. Snacking on whole almonds for 6 weeks improves endothelial function and lowers LDL cholesterol but does not affect liver fat and other cardiometabolic risk factors in healthy adults: the ATTIS study, a randomized controlled trial. Am J Clin Nutr 2020;111(6):1178-89. doi: 10.1093/ajcn/nqaa100.

7.	Domènech M, Serra-Mir M, Roth I, Freitas-Simoes T, Valls-Pedret C, Cofán M, López A, Sala-Vila A, Calvo C, Rajaram S, et al. Effect of a Walnut Diet on Office and 24-Hour Ambulatory Blood Pressure in Elderly Individuals. Hypertension 2019;73(5):1049-57. doi: 10.1161/hypertensionaha.118.12766.

8.	Estruch R, Martínez-González MA, Corella D, Salas-Salvadó J, Ruiz-Gutiérrez V, Covas MI, Fiol M, Gómez-Gracia E, López-Sabater MC, Vinyoles E, et al. Effects of a Mediterranean-style diet on cardiovascular risk factors: a randomized trial. Annals of internal medicine 2006;145(1):1-11. doi: 10.7326/0003-4819-145-1-200607040-00004.

9.	Fatahi S, Haghighatdoost F, Larijani B, Azadbakht L. Effect of Weight Reduction Diets Containing Fish, Walnut or Fish plus Walnut on Cardiovascular Risk Factors in Overweight and Obese Women. Arch Iran Med 2019;22(10):574-83.

10.	Huguenin GV, Moreira AS, Siant'Pierre TD, Gonçalves RA, Rosa G, Oliveira GM, Luiz RR, Tibirica E. Effects of Dietary Supplementation with Brazil Nuts on Microvascular Endothelial Function in Hypertensive and Dyslipidemic Patients: A Randomized Crossover Placebo-Controlled Trial. Microcirculation (New York, NY : 1994) 2015;22(8):687-99. doi: 10.1111/micc.12225.

11.	Hwang HJ, Liu Y. Daily walnut intake improves metabolic syndrome status and increases circulating adiponectin levels: randomized controlled crossover trial. 2019;13(2):105-14. doi: 10.4162/nrp.2019.13.2.105.

12.	Jenkins DJA, Kendall CWC, Lamarche B, Banach MS, Srichaikul K, Vidgen E, Mitchell S, Parker T, Nishi S, Bashyam B, et al. Nuts as a replacement for carbohydrates in the diabetic diet: a reanalysis of a randomised controlled trial. Diabetologia 2018;61(8):1734-47. doi: 10.1007/s00125-018-4628-9.

13.	Johnston CS, Sweazea KL, Schwab E, McElaney EA. Almond ingestion contributes to improved cardiovascular health in sedentary older adults participating in a walking intervention: A pilot study. Journal of Functional Foods 2017;39:58-62. doi: https://doi.org/10.1016/j.jff.2017.10.010.

14.	Jung H, Chen CO, Blumberg JB, Kwak HK. The effect of almonds on vitamin E status and cardiovascular risk factors in Korean adults: a randomized clinical trial. Eur J Nutr 2018;57(6):2069-79. doi: 10.1007/s00394-017-1480-5.

15.	Kasliwal RR, Bansal M, Mehrotra R, Yeptho KP, Trehan N. Effect of pistachio nut consumption on endothelial function and arterial stiffness. Nutrition (Burbank, Los Angeles County, Calif) 2015;31(5):678-85. doi: 10.1016/j.nut.2014.10.019.

16.	Li SC, Liu YH, Liu JF, Chang WH, Chen CM, Chen CY. Almond consumption improved glycemic control and lipid profiles in patients with type 2 diabetes mellitus. Metabolism 2011;60(4):474-9. doi: 10.1016/j.metabol.2010.04.009.

17.	Petrović-Oggiano G, Debeljak-Martačić J, Ranković S, Pokimica B, Mirić A, Glibetić M, Popović T. The Effect of Walnut Consumption on n-3 Fatty Acid Profile of Healthy People Living in a Non-Mediterranean West Balkan Country, a Small Scale Randomized Study. Nutrients 2020;12(1). doi: 10.3390/nu12010192.

18.	Richmond K, Williams S, Mann J, Brown R, Chisholm A. Markers of cardiovascular risk in postmenopausal women with type 2 diabetes are improved by the daily consumption of almonds or sunflower kernels: a feeding study. ISRN Nutr 2012;2013:626414-. doi: 10.5402/2013/626414.

19.	Ros E, Núñez I, Pérez-Heras A, Serra M, Gilabert R, Casals E, Deulofeu R. A walnut diet improves endothelial function in hypercholesterolemic subjects: a randomized crossover trial. Circulation 2004;109(13):1609-14. doi: 10.1161/01.cir.0000124477.91474.ff.

20.	Sanchis P, Molina M, Berga F, Muñoz E, Fortuny R, Costa-Bauzá A. A Pilot Randomized Crossover Trial Assessing the Safety and Short-Term Effects of Walnut Consumption by Patients with Chronic Kidney Disease. 2019;12(1). doi: 10.3390/nu12010063.

21.	Sari I, Baltaci Y, Bagci C, Davutoglu V, Erel O, Celik H, Ozer O, Aksoy N, Aksoy M. Effect of pistachio diet on lipid parameters, endothelial function, inflammation, and oxidative status: a prospective study. Nutrition (Burbank, Los Angeles County, Calif) 2010;26(4):399-404. doi: 10.1016/j.nut.2009.05.023.

22.	Sauder KA, McCrea CE, Ulbrecht JS, Kris-Etherton PM, West SG. Pistachio nut consumption modifies systemic hemodynamics, increases heart rate variability, and reduces ambulatory blood pressure in well-controlled type 2 diabetes: a randomized trial. Journal of the American Heart Association 2014;3(4). doi: 10.1161/jaha.114.000873.

23.	Schutte AE, Van Rooyen JM, Huisman HW, Mukuddem-Petersen J, Oosthuizen W, Hanekom SM, Jerling JC. Modulation of baroreflex sensitivity by walnuts versus cashew nuts in subjects with metabolic syndrome. Am J Hypertens 2006;19(6):629-36. doi: 10.1016/j.amjhyper.2005.12.014.

24.	Sheridan MJ, Cooper JN, Erario M, Cheifetz CE. Pistachio nut consumption and serum lipid levels. J Am Coll Nutr 2007;26(2):141-8. doi: 10.1080/07315724.2007.10719595.

25.	Sweazea KL, Johnston CS, Ricklefs KD, Petersen KN. Almond supplementation in the absence of dietary advice significantly reduces C-reactive protein in subjects with type 2 diabetes. Journal of Functional Foods 2014;10:252-9. doi: https://doi.org/10.1016/j.jff.2014.06.024.

26.	Williams PT, Bergeron N, Chiu S, Krauss RM. A randomized, controlled trial on the effects of almonds on lipoprotein response to a higher carbohydrate, lower fat diet in men and women with abdominal adiposity. Lipids in health and disease 2019;18(1):83. doi: 10.1186/s12944-019-1025-4.

27.	Agebratt C, Ström E, Romu T, Dahlqvist-Leinhard O, Borga M, Leandersson P, Nystrom FH. A randomized study of the effects of additional fruit and nuts consumption on hepatic fat content, cardiovascular risk factors and basal metabolic rate. PloS one 2016;11(1):e0147149.

28.	Aghasadeghi K, Nezhad MJZ, Hakimi H, Khazraei H. The effect of walnut oil consumption on blood sugar control in patients with type two diabetes. Pielęgniarstwo Chirurgiczne i Angiologiczne/Surgical and Vascular Nursing;2019(2):78-82.

29.	Alper CM, Mattes RD. Peanut consumption improves indices of cardiovascular disease risk in healthy adults. Journal of the American College of Nutrition 2003;22(2):133-41.

30.	Barceló F, Perona JS, Prades J, Funari SS, Gomez-Gracia E, Conde M, Estruch R, Ruiz-Gutiérrez V. Mediterranean-style diet effect on the structural properties of the erythrocyte cell membrane of hypertensive patients: the Prevencion con Dieta Mediterranea Study. Hypertension 2009;54(5):1143-50.

31.	Conlin PR, Chow D, Miller ER, Svetkey LP, Lin P-H, Harsha DW, Moore TJ, Sacks FM, Appel LJ. The effect of dietary patterns on blood pressure control in hypertensive patients: results from the Dietary Approaches to Stop Hypertension (DASH) trial. American journal of hypertension 2000;13(9):949-55.

32.	Costa e Silva LM, Pereira de Melo ML, Faro Reis FV, Monteiro MC, Dos Santos SM, Quadros Gomes BA, Meller da Silva LH. Comparison of the Effects of Brazil Nut Oil and Soybean Oil on the Cardiometabolic Parameters of Patients with Metabolic Syndrome: A Randomized Trial. Nutrients 2020;12(1):46.

33.	Davidi A, Reynolds J, Njike V, Ma Y, Doughty K, Katz D. The effect of the addition of daily fruit and nut bars to diet on weight, and cardiac risk profile, in overweight adults. Journal of Human Nutrition and Dietetics 2011;24(6):543-51.

34.	Davis CR, Bryan J, Hodgson JM, Woodman R, Murphy KJ. A Mediterranean diet reduces F2-isoprostanes and triglycerides among older Australian men and women after 6 months. The Journal of nutrition 2017;147(7):1348-55.

35.	Fatahi S, Haghighatdoost F, Larijani B, Azadbakht L. Effect of weight reduction diets containing fish, walnut or fish plus walnut on cardiovascular risk factors in overweight and obese women. Archives of Iranian medicine 2019;22(10):574-83.

36.	Ha AW, Kim WK, Kim JH, Kang NE. The supplementation effects of peanut sprout on reduction of abdominal fat and health indices in overweight and obese women. Nutr Res Pract 2015;9(3):249-55. doi: 10.4162/nrp.2015.9.3.249.

37.	Jenkins DJ, Kendall CW, Marchie A, Faulkner D, Vidgen E, Lapsley KG, Trautwein EA, Parker TL, Josse RG, Leiter LA. The effect of combining plant sterols, soy protein, viscous fibers, and almonds in treating hypercholesterolemia. Metabolism 2003;52(11):1478-83.

38.	Jenkins DJ, Kendall CW, Popovich DG, Vidgen E, Mehling CC, Vuksan V, Ransom TP, Rao AV, Rosenberg-Zand R, Tariq N. Effect of a very-high-fiber vegetable, fruit, and nut diet on serum lipids and colonic function. Metabolism-Clinical and Experimental 2001;50(4):494-503.

39.	Jones JB, Provost M, Keaver L, Breen C, Ludy M-J, Mattes RD. A randomized trial on the effects of flavorings on the health benefits of daily peanut consumption. The American journal of clinical nutrition 2014;99(3):490-6.

40.	Kaseb F, Rashidi M, Afkhami-Ardekani M, Fallahzadeh H. Effect of olive, almond and walnut oil on cardiovascular risk factors in type 2 diabetic patients. International Journal of Diabetes in Developing Countries 2013;33(2):115-9.

41.	Nasca MM, Zhou J-R, Welty FK. Effect of soy nuts on adhesion molecules and markers of inflammation in hypertensive and normotensive postmenopausal women. The American journal of cardiology 2008;102(1):84-6.

42.	Rabiei K, Ebrahimzadeh MA, Saeedi M, Bahar A, Akha O, Kashi Z. Effects of a hydroalcoholic extract of Juglans regia (walnut) leaves on blood glucose and major cardiovascular risk factors in type 2 diabetic patients: a double-blind, placebo-controlled clinical trial. BMC complementary and alternative medicine 2018;18(1):1-7.

43.	Razquin C, Martinez JA, Martinez‐Gonzalez MA, Fernández‐Crehuet J, Santos JM, Marti A. A Mediterranean diet rich in virgin olive oil may reverse the effects of the‐174G/C IL6 gene variant on 3‐year body weight change. Molecular nutrition & food research 2010;54(S1):S75-S82.

44.	Sanchez-Aguadero N, Garcia-Ortiz L, Patino-Alonso MC, Mora-Simon S, Gomez-Marcos MA, Alonso-Dominguez R, Sanchez-Salgado B, Recio-Rodriguez JI. Postprandial effect of breakfast glycaemic index on vascular function, glycaemic control and cognitive performance (BGI study): study protocol for a randomised crossover trial. Trials 2016;17(1):1-8.

45.	Storniolo CE, Casillas R, Bulló M, Castañer O, Ros E, Sáez G, Toledo E, Estruch R, Ruiz-Gutiérrez V, Fitó M. A Mediterranean diet supplemented with extra virgin olive oil or nuts improves endothelial markers involved in blood pressure control in hypertensive women. European journal of nutrition 2017;56(1):89-97.

46.	Tey SL, Robinson T, Gray AR, Chisholm AW, Brown RC. Do dry roasting, lightly salting nuts affect their cardioprotective properties and acceptability? 2017;56(3):1025-36. doi: 10.1007/s00394-015-1150-4.

47.	Tindall AM, McLimans CJ, Petersen KS, Kris-Etherton PM, Lamendella R. Walnuts and vegetable oils containing oleic acid differentially affect the gut microbiota and associations with cardiovascular risk factors: follow-up of a randomized, controlled, feeding trial in adults at risk for cardiovascular disease. The Journal of nutrition 2020;150(4):806-17.

48.	Toledo E, Hu FB, Estruch R, Buil-Cosiales P, Corella D, Salas-Salvadó J, Covas MI, Arós F, Gómez-Gracia E, Fiol M. Effect of the Mediterranean diet on blood pressure in the PREDIMED trial: results from a randomized controlled trial. BMC medicine 2013;11(1):1-10.

49.	Tovar J, Johansson M, Björck I. A multifunctional diet improves cardiometabolic-related biomarkers independently of weight changes: an 8-week randomized controlled intervention in healthy overweight and obese subjects. Eur J Nutr 2016;55(7):2295-306. doi: 10.1007/s00394-015-1039-2.

50.	Welty FK, Lee KS, Lew NS, Zhou JR. Effect of soy nuts on blood pressure and lipid levels in hypertensive, prehypertensive, and normotensive postmenopausal women. Archives of internal medicine 2007;167(10):1060-7. doi: 10.1001/archinte.167.10.1060.

51.	Wien MA, Sabaté JM, Iklé DN, Cole SE, Kandeel FR. Almonds vs complex carbohydrates in a weight reduction program. International journal of obesity and related metabolic disorders : journal of the International Association for the Study of Obesity 2003;27(11):1365-72. doi: 10.1038/sj.ijo.0802411.

52.	Zibaeenezhad M, Aghasadeghi K, Hakimi H, Yarmohammadi H, Nikaein F. The effect of walnut oil consumption on blood sugar in patients with diabetes mellitus type 2. International journal of endocrinology and metabolism 2016;14(3).

53.	Alves RD, Moreira AP, Macedo VS, de Cássia Gonçalves Alfenas R, Bressan J, Mattes R, Costa NM. Regular intake of high-oleic peanuts improves fat oxidation and body composition in overweight/obese men pursuing a energy-restricted diet. Obesity (Silver Spring, Md) 2014;22(6):1422-9. doi: 10.1002/oby.20746.

54.	Baer DJ, Gebauer SK. Walnuts Consumed by Healthy Adults Provide Less Available Energy than Predicted by the Atwater Factors. 2016;146(1):9-13. doi: 10.3945/jn.115.217372.

55.	Baer DJ, Novotny JA. Metabolizable Energy from Cashew Nuts is Less than that Predicted by Atwater Factors. Nutrients 2018;11(1):33. doi: 10.3390/nu11010033.

56.	Canales A, Benedi J, Bastida S, Corella D, Guillen M, Librelotto J, Nus M, Sánchez-Muniz FJ. The effect of consuming meat enriched in walnut paste on platelet aggregation and thrombogenesis varies in volunteers with different apolipoprotein A4 genotype. Nutricion hospitalaria 2010;25(5):746-54.

57.	Edwards K, Kwaw I, Matud J, Kurtz I. Effect of pistachio nuts on serum lipid levels in patients with moderate hypercholesterolemia. J Am Coll Nutr 1999;18(3):229-32. doi: 10.1080/07315724.1999.10718856.

58.	Hernández-Alonso P, Salas-Salvadó J, Baldrich-Mora M, Mallol R, Correig X, Bulló M. Effect of pistachio consumption on plasma lipoprotein subclasses in pre-diabetic subjects. Nutrition, metabolism, and cardiovascular diseases : NMCD 2015;25(4):396-402. doi: 10.1016/j.numecd.2015.01.013.

59.	Huguenin GV, Oliveira GM, Moreira AS, Saint'Pierre TD, Gonçalves RA, Pinheiro-Mulder AR, Teodoro AJ, Luiz RR, Rosa G. Improvement of antioxidant status after Brazil nut intake in hypertensive and dyslipidemic subjects. Nutr J 2015;14:54. doi: 10.1186/s12937-015-0043-y.

60.	Nishi SK, Kendall CW, Bazinet RP, Bashyam B, Ireland CA, Augustin LS, Blanco Mejia S, Sievenpiper JL, Jenkins DJ. Nut consumption, serum fatty acid profile and estimated coronary heart disease risk in type 2 diabetes. Nutrition, metabolism, and cardiovascular diseases : NMCD 2014;24(8):845-52. doi: 10.1016/j.numecd.2014.04.001.

61.	Salas-Huetos A, Muralidharan J, Galiè S, Salas-Salvadó J. Effect of Nut Consumption on Erectile and Sexual Function in Healthy Males: A Secondary Outcome Analysis of the FERTINUTS Randomized Controlled Trial. 2019;11(6). doi: 10.3390/nu11061372.

62.	Dikariyanto V, Smith L, Chowienczyk PJ, Berry SE, Hall WL. Snacking on Whole Almonds for Six Weeks Increases Heart Rate Variability during Mental Stress in Healthy Adults: A Randomized Controlled Trial. Nutrients 2020;12(6):1828.

63.	Domènech M, Serra-Mir M, Roth I, Freitas-Simoes T, Valls-Pedret C, Cofán M, López A, Sala-Vila A, Calvo C, Rajaram S. Effect of a walnut diet on office and 24-hour ambulatory blood pressure in elderly individuals: Findings from the WAHA randomized trial. Hypertension 2019;73(5):1049-57.

64.	Jamshed H, Sultan FAT, Amin F, Arslan J, Ghani S, Masroor M. Almond supplementation reduces serum uric acid in coronary artery disease patients: a randomized controlled trial. Nutrition journal 2015;15(1):1-5.

65.	Li S-C, Liu Y-H, Liu J-F, Chang W-H, Chen C-M, Chen C-YO. Almond consumption improved glycemic control and lipid profiles in patients with type 2 diabetes mellitus. Metabolism 2011;60(4):474-9.

66.	Martínez-González MA, Fernandez-Lazaro CI, Toledo E, Díaz-López A, Corella D, Goday A, Romaguera D, Vioque J, Alonso-Gómez ÁM, Wärnberg J. Carbohydrate quality changes and concurrent changes in cardiovascular risk factors: a longitudinal analysis in the PREDIMED-Plus randomized trial. The American journal of clinical nutrition 2020;111(2):291-306.

67.	Nishi S, Kendall CW, Gascoyne A-M, Bazinet RP, Bashyam B, Lapsley KG, Augustin LS, Sievenpiper JL, Jenkins DJ. Effect of almond consumption on the serum fatty acid profile: a dose–response study. British journal of nutrition 2014;112(7):1137-46.

68.	Schutte AE, Van Rooyen JM, Huisman HW, Mukuddem-Petersen J, Oosthuizen W, Hanekom SM, Jerling JC. Modulation of baroreflex sensitivity by walnuts versus cashew nuts in subjects with metabolic syndrome. American journal of hypertension 2006;19(6):629-36.

69.	Sureda A, Bibiloni MdM, Martorell M, Buil‐Cosiales P, Marti A, Pons A, Tur JA, Martinez‐Gonzalez MÁ, Investigators PS. Mediterranean diets supplemented with virgin olive oil and nuts enhance plasmatic antioxidant capabilities and decrease xanthine oxidase activity in people with metabolic syndrome: The PREDIMED study. Molecular nutrition & food research 2016;60(12):2654-64.

70.	Bhardwaj R, Dod H, Sandhu MS, Bedi R, Dod S, Konat G, Chopra HK, Sharma R, Jain AC, Nanda N. Acute effects of diets rich in almonds and walnuts on endothelial function. Indian heart journal 2018;70(4):497-501. doi: 10.1016/j.ihj.2018.01.030.

71.	Solà R, Valls RM, Godàs G, Perez-Busquets G, Ribalta J, Girona J, Heras M, Cabré A, Castro A, Domenech G. Cocoa, hazelnuts, sterols and soluble fiber cream reduces lipids and inflammation biomarkers in hypertensive patients: a randomized controlled trial. PloS one 2012;7(2):e31103.

72.	Wu H, Pan A, Yu Z, Qi Q, Lu L, Zhang G, Yu D, Zong G, Zhou Y, Chen X, et al. Lifestyle counseling and supplementation with flaxseed or walnuts influence the management of metabolic syndrome. The Journal of nutrition 2010;140(11):1937-42. doi: 10.3945/jn.110.126300.

73.	Gulati S, Misra A, Pandey RM. Effect of Almond Supplementation on Glycemia and Cardiovascular Risk Factors in Asian Indians in North India with Type 2 Diabetes Mellitus: A 24-Week Study. Metabolic syndrome and related disorders 2017;15(2):98-105. doi: 10.1089/met.2016.0066.

74.	Michels AJ, Leonard SW, Uesugi SL, Bobe G, Frei B, Traber MG. Daily Consumption of Oregon Hazelnuts Affects α-Tocopherol Status in Healthy Older Adults: A Pre-Post Intervention Study. J Nutr 2018;148(12):1924-30. doi: 10.1093/jn/nxy210.

75.	Barbour JA, Howe PRC. Cerebrovascular and cognitive benefits of high-oleic peanut consumption in healthy overweight middle-aged adults. 2017;20(10):555-62. doi: 10.1080/1028415x.2016.1204744.

76.	Choudhury K, Clark J, Griffiths HR. An almond-enriched diet increases plasma α-tocopherol and improves vascular function but does not affect oxidative stress markers or lipid levels. Free radical research 2014;48(5):599-606. doi: 10.3109/10715762.2014.896458.
